# Real-World Practices of Fluoroquinolone Prophylaxis in Spontaneous Bacterial Peritonitis: A Longitudinal Study from a Tertiary Care Center in North India

**DOI:** 10.64898/2026.07.14.26357717

**Authors:** Anika Malviya, Prasan Kumar Panda, Anand Sharma, Ravi Kant, Mukesh Bairwa, Vikas Panwar, Bhupinder Solanki, Ruchi Dua

## Abstract

**Background and objectives:** Spontaneous bacterial peritonitis (SBP) is a life-threatening complication of cirrhosis with ascites, carrying one- and two-year mortality rates exceeding 70% and 80%, respectively. Fluoroquinolone prophylaxis is the cornerstone of SBP prevention. Real-world longitudinal data on prescribing practices and clinical outcomes from Indian tertiary care centers are sparse. We aimed to evaluate fluoroquinolone prescribing patterns, guideline adherence, and six-month clinical outcomes in SBP patients at a tertiary academic center in North India.

**Methods:** This was a pre-specified sub-analysis of a 15-month analytical longitudinal study at AIIMS Rishikesh. Adults (age ≥18 years) admitted with SBP and initiated on fluoroquinolone prophylaxis were enrolled consecutively and followed for six months. Prescribing practices were compared against EASL and AASLD recommendations. The primary outcome was the rate of guideline-directed prescribing. Secondary outcomes included clinical cure at discharge, six-month cure, relapse, regimen modification, adverse drug reactions, and treatment compliance. Categorical variables were compared by Fisher’s exact test or chi-squared test (SPSS).

**Results:** Forty-eight SBP patients were included (mean age 44.75 ± 11.94 years; 85.4% male). Guideline-directed fluoroquinolone prophylaxis was prescribed to all patients (100%). Norfloxacin 400 mg once daily was predominant (85.4%), followed by levofloxacin (10.4%) and moxifloxacin (4.2%). Cure at discharge was 85.4%. At six months, 64.6% maintained sustained cure and 22.9% relapsed. Regimen modification occurred in 22.9%, most commonly antimicrobial substitution. Nausea was the only adverse drug reaction (4.8%). Treatment compliance was 73.8%. No patient underwent therapeutic drug monitoring.

**Conclusions:** Fluoroquinolone prescribing for SBP prophylaxis at AIIMS Rishikesh was fully concordant with standard guidelines. Despite complete adherence, a relapse rate of 22.9% and frequent regimen modification underscore the limitations of long-term fluoroquinolone prophylaxis, likely reflecting emerging quinolone resistance. Strengthening antimicrobial stewardship is essential to sustain prophylaxis effectiveness in Indian tertiary care settings.

**Trial registration:** CTRI/2025/06/088655

## Introduction

Spontaneous bacterial peritonitis (SBP) is a frequent and often fatal complication of cirrhosis with ascites, occurring in 7–30% of hospitalised cirrhotic patients (1,2). Mortality from a single episode, even with appropriate treatment, approaches 20–40%, and one-year and two-year mortality rates following a first SBP episode have been reported to exceed 70% and 80%, respectively, reflecting the severity of the underlying liver disease rather than the infection alone (3). Without secondary prophylaxis, recurrence occurs in approximately 70% of patients within one year (4).

Fluoroquinolones, particularly norfloxacin, are the standard agents for both primary and secondary SBP prophylaxis. Both the EASL and the AASLD recommend norfloxacin 400 mg orally once daily, continued until liver transplantation or death, in patients who have survived a prior SBP episode (5,6). The pharmacodynamic rationale for this regimen rests on the concentration-dependent bactericidal activity and prolonged post-antibiotic effect of fluoroquinolones, which allows once-daily dosing to achieve effective gut decontamination against gram-negative Enterobacteriaceae – the principal organisms implicated in SBP pathogenesis through intestinal bacterial translocation (7,8).

Despite guideline recommendations, the widespread and prolonged use of fluoroquinolones in cirrhotic patients has raised concerns about the emergence of quinolone-resistant organisms, particularly in South Asian settings where antibiotic stewardship programmes are still evolving (9,10). Several studies from India as well as from other countries have documented increasing fluoroquinolone resistance among gram-negative organisms causing SBP, which may reduce the effectiveness of standard prophylaxis regimens over time (11,12).

Despite the high prevalence of cirrhosis and its complications in India, longitudinal observational data specifically evaluating fluoroquinolone prescribing practices, adherence to published guidelines, and downstream clinical outcomes for SBP prophylaxis from Indian tertiary care centers are limited. In India, antimicrobial stewardship capabilities are often described as underdeveloped or trivial, resulting in a significant lack of empirical evidence and insufficient documentation regarding compliance with guidelines (13). This study addresses that gap.

We report a longitudinal sub-analysis of fluoroquinolone prescribing practices and six-month clinical outcomes in patients admitted with SBP to AIIMS Rishikesh, a high-volume tertiary academic medical center serving a large referral population in North India. Our aim was to characterize prescribing patterns, determine guideline adherence rates, and evaluate outcomes at 6 months including relapse, adverse drug reactions, and treatment compliance.

## Methods

### Study design and setting

This is a pre-specified sub-analysis of a 15-month (February 2025 - May 2026) analytical longitudinal observational study conducted in the Gastroenterology and Internal Medicine departments of All India Institute of Medical Sciences (AIIMS) Rishikesh, Uttarakhand, India. AIIMS Rishikesh is a central government tertiary care academic institution that serves as the primary referral center for North India, primarily receiving patients from the states of Uttarakhand and Uttar Pradesh.

### Study population

Consecutive patients aged 18 years or above, admitted with a diagnosis of SBP, and initiated on fluoroquinolone prophylaxis who remained hospitalized for more than 48 hours were enrolled. SBP was diagnosed on the basis of an ascitic fluid polymorphonuclear (PMN) cell count ≥250 cells/mm^3^, with or without a positive ascitic fluid culture, in the absence of a surgically treatable source of peritoneal infection. Patients discharged within 48 hours of admission and those who did not complete the prescribed duration of therapy were excluded.

### Data collection

Data were collected using a standardised electronic proforma and finalized in Microsoft Excel. Recorded variables included: demographic information, comorbidities, clinical presentation, diagnosis and diagnostic basis, antimicrobial agent and dose, dosing frequency and schedule, type of therapy (prophylactic, empirical, or definitive), and guideline reference cited by the treating team. Fluoroquinolone regimens studied included norfloxacin, levofloxacin, ciprofloxacin, and moxifloxacin when used for SBP prophylaxis or treatment.

### Definitions

Guideline adherence was defined as prescription of norfloxacin 400 mg once daily, or an equivalent regimen, in accordance with EASL and AASLD recommendations [4,5]. Clinical cure at discharge was defined as resolution of signs and symptoms of SBP with clinical improvement adequate for hospital discharge. Sustained cure at six months was defined as the absence of SBP relapse at the six-month follow-up assessment. Relapse was defined as a new microbiologically or clinically confirmed episode of SBP during the six-month follow-up period. Treatment compliance was defined as intake of 80% or more of prescribed doses during follow-up, as reported by the patient at telephonic contact.

### Follow-up

All enrolled participants were followed for six months after hospital discharge. Follow-up data were collected through outpatient clinic records and telephonic interview at the six-month time point. Information on SBP relapse, treatment compliance, adverse events, and regimen changes was documented.

### Ethical approval

The study was approved by the Institutional Ethics Committee of AIIMS Rishikesh (reference number AIIMS/IEC/25/178, dated 28 February 2025). The study was prospectively registered in the Clinical Trials Registry of India (CTRI/2025/06/088655). All procedures were conducted in accordance with the Declaration of Helsinki as revised in 2013. Written informed consent was obtained from every participant prior to enrollment.

### Statistical analysis

Data were analyzed using SPSS software. Continuous variables following normal distribution were expressed as mean ± standard deviation (SD); non-normally distributed variables were expressed as median with interquartile range (IQR). Categorical variables were expressed as frequencies and percentages with 95% confidence intervals (CI). Associations between categorical variables were assessed using the chi-squared test or Fisher’s exact test as appropriate. A two-tailed *p* value <0.05 was considered statistically significant throughout.

## Results

### Patient enrollment and baseline characteristics

Of 296 patients screened across four disease groups in the parent study, 48 patients with SBP who received fluoroquinolone prophylaxis were eligible and included in this analysis. The mean age was 44.75 ± 11.94 years, with the majority (85.4%, n = 41) being male. This demographic profile reflects the well-established male preponderance of alcohol-related chronic liver disease in North India (14). The mean duration of hospital stay was 10.21 ± 5.73 days (range 3–54 days). Ninety-three point eight percent of infections were community-acquired (45/48). All patients had chronic liver disease as the primary underlying diagnosis. Baseline characteristics are summarized in Table 1.

**Table 1.** Baseline characteristics of the SBP cohort (n = 48).

| Parameter | Value |
| --- | --- |
| Age (years), mean ± SD | 44.75 ± 11.94 |
| Gender, n (M:F) | 41 : 7 (85.4% M) |
| Length of hospital stay (days), mean ± SD | 10.21 ± 5.73 |
| Range of length of hospital stay (days) | 3 – 54 |
| Community-acquired infection, n (%) | 45 (93.8%) |
| Hospital-acquired infection, n (%) | 3 (6.3%) |
| Prophylaxis duration prescribed (days), mean ± SD | 180.0 ± 0.0 |

### Fluoroquinolone prescribing patterns

All 48 patients received fluoroquinolone prophylaxis. Norfloxacin 400 mg once daily was the predominant regimen (n = 41; 85.4%), followed by levofloxacin (750 mg in 4 patients and 500 mg in 1 patient) (n = 5; 10.4%) and moxifloxacin 400 mg (n = 2; 4.2%). Among levofloxacin recipients, once-daily dosing was used in four patients and alternate-day dosing in one. Both moxifloxacin prescriptions were once daily. Ciprofloxacin was not used in this cohort for SBP prophylaxis. All SBP patients were prescribed prophylactic therapy with a mean (± SD) duration of 180.0 ± 0.0 days, indicating continuation of prophylaxis throughout the entire six-month follow-up period in all cases. EASL and AASLD guidelines were the exclusive references cited for all SBP prescriptions. Prescribing details are shown in Table 2.

**Table 2.** Fluoroquinolone prescribing patterns in the SBP cohort (n = 48).

| Agent | n (%) | Dosing schedule | Guideline |
| --- | --- | --- | --- |
| Norfloxacin 400 mg | 41 (85.4%) | Once daily | EASL / AASLD |
| Levofloxacin 750 mg (n = 4)<br>Levofloxacin 500 mg (n=1) | 5 (10.4%) | Once daily (n = 4);<br>alternate day 750 mg<br>(n = 1) | EASL |
| Moxifloxacin 400 mg | 2 (4.2%) | Once daily | EASL |

EASL European Association for the Study of the Liver; AASLD American Association for the Study of Liver Diseases.

### Guideline adherence and regimen modifications

Guideline-directed dosing was observed in all 48 patients (100%), representing the highest adherence rate among the four diagnostic groups in the parent study (Fisher’s exact test, *p* = 0.018). In contrast, the recurrent urinary tract infection, bronchiectasis, and cryptococcal meningitis subgroups had physician-determined dosing in 6.2%, 16.0%, and 12.5% of cases, respectively. Regimen change over six months was required in 11 patients (22.9%), with a median time to change of 60 (IQR 30–90) days; 68.2% of regimen changes occurred within the first two months. The most common type of modification was substitution with a new antimicrobial (n = 9; 81.8%), while two patients had treatment discontinued (18.2%). No patient in the SBP cohort received therapeutic drug monitoring, dosing nomogram guidance, or software-assisted dose optimization during the six-month follow-up. Data are presented in Table 3.

**Table 3.**
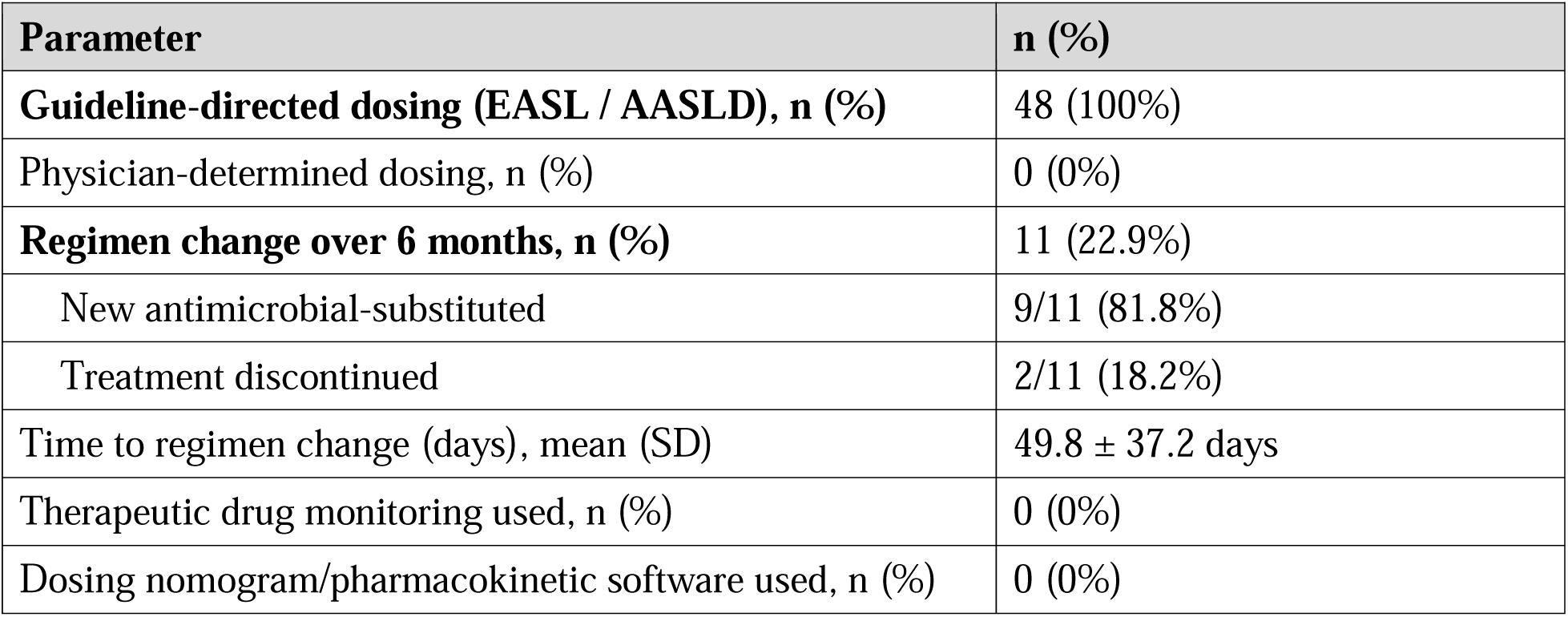
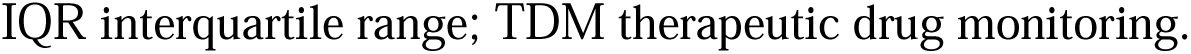
Guideline adherence and regimen modification summary (n = 48).

### Clinical outcomes

Cure at discharge was achieved in 41 patients (85.4%; 95% CI: 72.2–93.9%), with five (10.4%) showing clinical improvement and two (4.2%) classified as not cured. At six-month follow-up, sustained cure was maintained in 31 patients (64.6%). Eleven patients (22.9%) experienced SBP relapse, three (6.3%) were lost to follow-up, and three (6.3%) died from causes unrelated to SBP. No statistically significant association was observed between source of dosing strategy and cure at discharge (*p* = 0.707), cure at six months (*p* = 1.000), or relapse at six months (*p* = 1.000) in the overall study. Treatment compliance was documented in 31 of 42 patients with complete follow-up data (73.8%; 95% CI: 58.0–86.1%), which was significantly lower than in the other three diagnostic groups (Fisher’s exact test, *p* = 0.001). Clinical outcomes are summarized in Table 4.

**Table 4.** Clinical outcomes at discharge and at six-month follow-up (n = 48).

| **Outcome** | **n (%)** | **95% CI** |
| --- | --- | --- |
| **Cure at discharge** | 41 (85.4%) | 72.2% – 93.9% |
| Improved at discharge | 5 (10.4%) | 3.5% – 23.1% |
| Not cured at discharge | 2 (4.2%) | 0.5% – 14.3% |
| **Sustained cure at 6 months** | 31 (64.6%) | 49.5% – 77.8% |
| Relapse at 6 months | 11 (22.9%) | 12.0% – 37.3% |
| Lost to follow-up | 3 (6.3%) | 1.3% – 17.2% |
| Death from other cause | 3 (6.3%) | 1.3% – 17.2% |
| **Treatment compliance ($\geq 80\%$ doses taken)** | 31/42†<br>(73.8%) | 58.0% – 86.1% |
| Adverse drug reaction – nausea only, n (%) | 2 (4.8%) | 0.6% – 16.2% |

†Compliance data available for 42 patients who completed six-month follow-up. CI confidence interval.

### Adverse drug reactions

The safety profile of fluoroquinolone prophylaxis was favorable overall. No adverse drug reactions were documented in 40 of 42 patients with ADR data available (95.2%). Nausea was the only drug-related adverse event, occurring in two patients (4.8%) receiving norfloxacin. No cases of tendinopathy, QT prolongation, hepatotoxicity, photosensitivity, or serious adverse events attributable to fluoroquinolones were recorded. Notably, no patient in the SBP cohort developed acute kidney injury. The association between antimicrobial agent and ADR type across the full study cohort was statistically significant (*p* = 0.017), confirming drug-specific rather than random ADR distribution.

## Discussion

The main finding of this longitudinal study is that fluoroquinolone prescribing for SBP prophylaxis at AIIMS Rishikesh was fully guideline-concordant (100%), with norfloxacin 400 mg once daily as the near-universal choice. Short-term outcomes were favourable, with 85.4% of patients achieving cure at discharge. However, a six-month relapse rate of 22.9% and the need for regimen modification in nearly one-quarter of patients highlight the inherent limitations of prolonged fluoroquinolone prophylaxis in a setting where quinolone resistance is an escalating concern.

Both EASL and AASLD guidelines specifically recommend norfloxacin 400 mg once daily as the first-line agent for long-term SBP prophylaxis (5,6). Norfloxacin achieves selective gut decontamination by targeting aerobic gram-negative enteric bacteria with minimal effect on anaerobic flora, thereby reducing the principal mechanism of SBP – bacterial translocation from the intestinal lumen to the mesenteric lymphatics and then to the peritoneal fluid (7,15). The once-daily regimen is pharmacodynamically justified by norfloxacin’s concentration-dependent bactericidal profile and its prolonged post-antibiotic effect (16,17). The predominance of norfloxacin in our cohort (85.4%) is directly aligned with these recommendations. The use of levofloxacin in 10.4% and moxifloxacin in 4.2% was due to the concomitant presence of tuberculosis and the patient receiving modified anti-tubercular therapy.

Compared with data from analogous tertiary centers, our guideline adherence rates are notably high. A pilot stewardship program at AIIMS New Delhi reported baseline guideline adherence of only 66% across all antimicrobial indications before intervention (18). A WHO methodology-based point prevalence survey at a major Pakistani tertiary hospital found that 61.9% of antimicrobials were prescribed without any guideline reference (19). In regional intensive care unit studies from India, empirical antibiotic use without microbiological confirmation approached 92% (20). The consistently high adherence observed in our SBP cohort is plausible given the clarity, international visibility, and direct clinical relevance of EASL and AASLD SBP guidelines, combined with structured faculty-supervised prescribing practices in academic departments.

The six-month relapse rate of 22.9% observed in this study, despite complete guideline adherence, is clinically important and consistent with published data from India and other countries with high fluoroquinolone use. Prolonged norfloxacin prophylaxis has been shown to gradually shift the microbiological profile of SBP toward gram-positive organisms and quinolone-resistant gram-negative strains, thereby reducing the long-term protective efficacy of fluoroquinolone-based regimens (11,21). The high proportion of antimicrobial substitutions among patients requiring regimen change (81.8%) in our cohort is consistent with this pattern of emerging resistance resulting in treatment failure.

Treatment compliance at 73.8% in the SBP group was significantly lower than in the other three diagnostic groups in the parent study (*p* = 0.001). This is not unexpected, given the requirement for life-long fluoroquinolone prophylaxis in a population with debilitating chronic liver disease, recurrent hospitalizations, and high out-of-pocket medication expenditure. In India, where patients directly bear a substantial proportion of healthcare costs, adherence to long-term oral prophylaxis is a recognized and persistent challenge (22,23). Incomplete compliance with fluoroquinolone prophylaxis may itself contribute to resistance selection by repeatedly exposing gut flora to sub-inhibitory drug concentrations, paradoxically worsening the problem it is intended to prevent.

The complete absence of therapeutic drug monitoring, pharmacokinetic-guided dosing, and dose-optimization software across all 48 SBP patients represents an important stewardship gap. Although TDM is not routinely indicated for fluoroquinolones in standard clinical practice, its absence for potentially nephrotoxic agents co-prescribed in patients with cirrhosis and renal impairment deserves attention. Development of institutional protocols for PK/PD-guided monitoring in this high-risk group warrants consideration.

This study has several important strengths. Its longitudinal design with six-month prospective follow-up provides a level of clinical detail that is rarely available in the existing Indian literature on SBP management. Data were collected from a well-characterized consecutive cohort at a high-volume tertiary academic center, making the findings clinically representative of this care setting. The study evaluated multiple outcome dimensions including relapse, ADR occurrence, regimen modifications, and treatment compliance in a single prospective cohort.

Limitations include the single-center design, which may limit generalizability to settings with different demographics and formulary access. The SBP subgroup size of 48 patients, while adequate for descriptive analysis, constrains the statistical power of association analyses. Microbiological culture data with susceptibility testing were not consistently available for all patients, precluding systematic assessment of pathogen distribution and resistance profiles. Finally, the complete absence of TDM precluded any objective pharmacokinetic-outcome analysis.

Future studies should prospectively incorporate systematic microbiological surveillance with fluoroquinolone susceptibility testing in patients receiving SBP prophylaxis to track resistance trends over time. Head-to-head comparative studies evaluating rifaximin – a non-absorbable antibiotic proposed as a less resistance-prone alternative for gut decontamination – against norfloxacin in the Indian population are needed (24,25). Integrating pharmacokinetic monitoring and structured adherence support programs into SBP prophylaxis protocols may further improve outcomes in this high-risk population.

## Conclusion

Fluoroquinolone prescribing for SBP prophylaxis at AIIMS Rishikesh was fully concordant with EASL and AASLD guidelines, with norfloxacin 400 mg once daily as the standard regimen. Short-term clinical outcomes were favorable, with cure achieved in 85.4% of patients at discharge. The six-month relapse rate of 22.9% and the requirement for regimen modification in 22.9% of patients highlight the limitations of prolonged fluoroquinolone prophylaxis, likely reflecting early emergence of quinolone resistance. The absence of therapeutic drug monitoring and lower treatment compliance in this group identify specific targets for antimicrobial stewardship interventions. Prospective studies with microbiological surveillance are warranted to track resistance trends and evaluate alternative prophylactic strategies in Indian tertiary care centers.

## Data Availability

All data produced in the present study are available upon reasonable request to the authors.

## Declarations

### Ethics approval

This study was performed in line with the principles of the Declaration of Helsinki. Approval was granted by the Institutional Ethics Committee of AIIMS Rishikesh (No. AIIMS/IEC/25/178, dated 28 February 2025). The study was registered in the Clinical Trials Registry of India (CTRI/2025/06/088655).

### Consent to participate

Written informed consent was obtained from all individual participants included in the study.

### Consent to publish

Not applicable (no identifying personal data or images are included).

### Data availability

The datasets generated and analyzed during the current study are available from the corresponding author on reasonable request.

